# Exploring the short-term role of particulate matter in the COVID-19 outbreak in USA cities

**DOI:** 10.1101/2021.03.09.21253212

**Authors:** Leonardo Yoshiaki Kamigauti, Gabriel Martins Palma Perez, Carlos Eduardo Souto-Oliveira, Elizabeth Cowdery, Paulo Hilário Nascimento Saldiva, Maria de Fatima Andrade

**Affiliations:** Department of Atmospheric Sciences, University of São Paulo, São Paulo, Brazil; Department of Meteorology, University of Reading, Earley Gate, United Kingdom; Department of Earth and Environment, University of Boston, Boston, United States of America; Department of Pathology, University of São Paulo, São Paulo, Brazil

**Author notes:** Corresponding author Email address (Leonardo Yoshiaki Kamigauti).

**Keywords:** COVID-19, SARS-CoV-2, airborne transmission, particulate matter, Granger’s Causality

## Abstract

The role of particulate matter (PM) in the COVID-19 pandemic is currently being discussed by the scientific community. Long-term (years) exposure to PM is known to affect human health by increasing susceptibility to viral infections as well as to the development of respiratory and cardiovascular symptoms. In the short-term (days to months), PM has been suggested to assist airborne viral transmission. However, confounding factors such as urban mobility prevent causal conclusions. In this study, we explore short-term relationships between PM concentrations and the evolution of COVID-19 cases in a number of cities in the United States of America. We focus on the role of PM in facilitating viral transmission in early stages of the pandemic. We analyzed PM concentrations in two particle size ranges, *<* 2.5 *µm*, and between 10 and 2.5 *µm* (PM_2.5_ and PM_10_ respectively) as well as carbon monoxide (CO) and nitrogen dioxide (NO_2_). Granger causality analysis was employed to identify instantaneous and lagged effects of pollution in peaks of COVID-19 new daily cases in each location. The effect of pollution in shaping the disease spread was evaluated by correlating the logistic growth rate of accumulated cases with pollutants concentrations for a range of time lags and accumulation windows. PM_2.5_ shows the most significant results in Granger causality tests in comparison with the other pollutants. We found a strong and significant association between PM_2.5_ concentrations and the growth rate of accumulated cases between the 1^*st*^ and 18^*th*^ days after the report of the infection, peaking at the 8^*th*^ day. By comparing results of PM_2.5_ with PM_10_, CO and NO_2_ we rule out confounding effects associated with mobility. We conclude that PM_2.5_ is not a first order effect in the cities considered; however, it plays a significant role in facilitating the COVID-19 transmission. We estimate that the growth rate of COVID-19 cases would be risen by 12.5% if PM_2.5_ is increased from 25 to 35 *µ*g *m*^−3^.

## 1. Introduction

The current pandemic scenario has encouraged the scientific community to gather resources in the characterization of the evolution of COVID-19 [1, 2]. One of the primary and most pressing concern to worldwide policy makers is the saturation of health systems caused by a high number of infections, as quantified by the reported number of cases per day (daily cases) [3]. Changes in daily cases are characterized by changes in the transmission rate of SARS-CoV-2, which is associated with a number of societal, environmental, and behavioral factors [4]. In this study, we explore the role of pollutants, namely airborne particulate matter (PM), carbon monoxide (CO) and nitrogen dioxide (NO_2_), on the evolution of the COVID-19 pandemics across the United States of America (USA).

Statistical links between air quality and COVID-19 daily cases are expected to occur through different mechanisms; in this study, we explore the following hypotheses: 1) PM has been suggested to facilitate viruses transmission, due to its role in shielding the virus during airborne transport [5, 6, 7, 8, 9]; 2) The long-term exposure, particularly to CO and NO_2_, increases the susceptibility of a population to adverse health conditions [10, 11]; 3) PM levels are associated with urban mobility [12], thus we expect them to respond to the effectiveness of local social distancing measures. In the following subsections we discuss in detail these mechanisms as well as the statistical techniques employed to explore them.

### 1.1. Aerosol-assisted SARS-CoV-2 spread

Short-term interactions between aerosol and airborne viruses can occur through a spreading assistance mechanism. This mechanism is dependent on the aerosol size. Large (*>* 5*µm*) and small droplets (*<* 5*µm*) containing viruses are released by infected persons when talking, coughing, sneezing or vomiting. While larger droplets travel shorter distances (1 – 2 meters), smaller droplets can travel up to tens of meters. In addition, suspended droplets can lose mass by drying and can also interact with other particles, physically and chemically, further changing their travel distances [9].

A growing body of research is investigating virus transport and transmission associated with PM. In Spain, deposition rates of billions of viruses per *m*^2^ per day were estimated considering long-range transport from air masses coming from marine and desert sources [13]. In China, influenza-like-illnesses were associated with PM with aerodynamic diameter lesser than 2.5 *µm* (PM_2.5_) in lags of 2 days during flu-season [8]. Chen et al. estimated that approximately 10% of influenza cases result from exposure to ambient PM_2.5_, suggesting that the reduction of PM_2.5_ concentrations lowers influenza transmission [7].

The preliminary evidence of SARS-CoV-2 interacting with PM was found at an industrial site in Bergamo Province, Italy. Samples detected viral RNA in PM [5]. Bergamo is characterized by high concentrations of PM and was severely affected by COVID-19. In addition, other studies have found viral RNA of SARS-CoV-2 in aerosol from hospitals and stores in Wuhan, China [14] and Nebraska, USA [2]. In the USA, researchers have shown that viruses survive in aerosol for at least 3 hours [15]. Tung et al. [16] present a review of studies about the virus-PM interaction.

Virus viability in PM is favored by key-compounds which contribute to its protection. Aerosol particles contain organic matrices of exopolymeric compounds, which absorb ultraviolet wavelengths and prevent the dehydration of the virus [17, 18]. In addition, viruses can be stabilized and protected from environmental stress and antimicrobial agents by colloidal and solid organic matter, such as biofilm [19]. A virus-PM interaction simulation [9] has shown enhancement of respiratory syncytial virus viability potentially due to the formation of a salt and carbon protective encasing; when associated with aerosol particles, the viruses remain capable of infection at low temperatures up to 6 months. In addition to providing protection, a virus-PM interaction has the potential to modify virus infectivity characteristics. Due to PM black carbon fraction, virus-PM interactions have been shown to accelerate virus entry and bioavailability in the cell [9, 20, 6].

### 1.2. Sources and Long-term health impacts of air pollution

The size and composition of PM is dependent on its source. Primary PM is ones produced directly in the source, whilst secondary PM is produced while in the atmosphere by gas-particle conversion processes. Vehicular (exhaust, tyre and break wear), soil dust ressuspension, and industrial emissions are the main sources of PM with aerodynamic diameters between 2.5 and 10 *µm* (PM_10_) in US urban centers [21, 22, 23, 24]. Secondary PM explains much of the mass of PM_2.5_, followed by fuel combustion [25, 21]. The source apportionment of primary PM is more accurate as it retains the source composition at some level. Secondary PM is mostly produced by non-linear reactions, thus, information about source characteristics is lost [26, 27].

The effects of PM exposure in human health, especially in long-term, is well know and discussed [28, 29, 30, 31]. The penetrability of PM in the respiratory system depends on the particle size. Finer particles have a greater penetration potential [32]. PM_10_ can penetrate the respiratory system mostly accumulating at the tracheobronchial tree. PM_2.5_ can reach the bronchioles and alveoli and penetrate the circulatory system [33]. PM concentration is also directly linked to viral infection [34]. Yao et al. suggest that long-term exposition to PM affects COVID-19 prognosis [35]. Thus, its very likely the PM exposure is directly linked to the COVID-19 symptoms development and the daily cases report.

Fossil fuel usage is the main source of CO and NO_2_ [36, 37, 38]. CO is relatively stable in the atmosphere with residence time of at least one month [39]. NO_2_ is involved in many atmospheric reactions (e.g. ozone formation and secondary aerosol formation). Therefore, its concentration is not linearly proportional to fossil fuel usage [27].

The effects of long-term exposure to CO and NO_2_ are matter of concern in many studies. The toxicity of CO is directly linked to cellular apoxia [40]. Short and long-term studies link chronicle exposure to CO with cardiovascular events and death [41]. There is no major link between ambient level CO and pulmonary complications [41]. NO_2_ is a strong oxidant that affects mainly the lungs, causing diverse respiratory symptoms; it also increases the susceptibility to viral infection [42]. The effect of long-term exposition of NO_2_ on mortality is comparable to PM_2.5_ exposition [43].

### 1.3. Statistical modelling and time series analysis

The incubation time of COVID-19 ranges from 4 to 15 days. Therefore, perturbations in the epidemic evolution caused by air pollution through airborne transmission would be observed in time lags within this same interval. It is simpler, however, to quantify the ability of air pollution to predict perturbations in the epidemic growth rather than asserting true causality. This can be done by computing linear correlations between the target variable (i.e., perturbations in cases) and the lagged explanatory variables (i.e., concentration of pollutants). In the econometric framework of Granger causality [44], it is possible to test if an explanatory variable, up to a number of lags in the past, contains useful information to forecast the target variable. If an explanatory variable pass this test above a significance threshold, it is said that it “Granger causes” the target variable.

Granger causality was employed in a number of studies related to air pollution. Jiang and Bai [45] investigated causal relationships between emissions in Beijing and its neighboring cities. Zhu et al. [46] identified elements of the urban dynamics that Granger cause air pollution in China. Wang et al. [47] employed Granger causality to relate mortality rates and air pollution. Mele and Magazzino [48] employed the same framework to investigate causal relationships between economic growth and air pollution in India.

More recently, studies investigating causes of new cases of COVID-19 and consequences of the global pandemic have employed the Granger framework. Bushman et al. [49] investigated the effect of social distancing on the number of deaths related to COVID-19. Awasthi et al. [50] inferred from causal models that temperature and humidity do not cause new cases of COVID-19.

The effect of air pollution on peaks of daily cases can be quantified by lagged correlations and Granger analysis. However, we also expect a relationship between the shape of the accumulated cases curve and pollution. In other words, we expect that the SARS-CoV-2 spread is faster in more polluted cities. To this end, we employed a logistic regression to extract shape parameters of the accumulated cases curve. The logistic regression has been shown to fit well COVID-19 cases in Italy and China [51, 52]. The analysis of the impact of PM2.5 as a carrier for the virus was performed to the EUA for two reasons, the high number of infected per day and the data accessibility to Pollutants concentrations and cases of COVID at many different cities and counties of the country.

## 2. Methodology

### 2.1. Data

The following datasets were employed in this study:

- PM and pollutant gases data were retrieved from the World Air Quality Index (WAQI) project at https://aqicn.org/data-platform/covid19 (September 2020). The WAQI dataset is available at city level in the USA.
- 2019 Novel Coronavirus COVID-19 (2019-nCoV) Data Repository by Johns Hopkins University Center for Systems Science and Engineering for COVID-19 data, available at https://github.com/CSSEGISandData/COVID-19 (September 2020). The data was retrieved at county level.

The period covered ranges from December 30, 2019 to July 31, 2020. Details in Table 1. WAQI aggregates data from local official monitoring datasets. City level pollution data were associated with the respective county level COVID-19 data by matching the metadata available in both datasets. Although ideally both datasets should be at the same administrative level, we expect this association is appropriated for this study since the cities in the WAQI dataset are the most populous of their respective counties.

**Table 1:** Dataset summary with descriptive statistics and missing data info. Note that this dataset is the one after the quality check described.

| Variable | Stats / Values | Freqs (% of Valid) | Valid | Missing |
| --- | --- | --- | --- | --- |
| County | 1. Albuquerque | 198 ( 1.9%) | 10296 | 0 |
|  | 2. Atlanta | 198 ( 1.9%) | (100.0%) | (0.0%) |
|  | 3. Austin | 198 ( 1.9%) |  |  |
|  | 4. Baltimore | 198 ( 1.9%) |  |  |
|  | 5. Boise | 198 ( 1.9%) |  |  |
|  | [ 47 others ] | 9306 (85.3%) |  |  |
| State | 1. Arizona | 396 ( 3.8%) | 10296 | 0 |
|  | 2. Arkansas | 198 ( 1.9%) | (100.0%) | (0.0%) |
|  | 3. California | 1188 (11.5%) |  |  |
|  | 4. Colorado | 198 ( 1.9%) |  |  |
|  | 5. Connecticut | 198 ( 1.9%) |  |  |
|  | [ 26 others ] | 8118 (78.8%) |  |  |
| Day | min : 2019-12-30 | 198 distinct values | 10296 | 0 |
|  | med : 2020-04-06 12:00:00 |  | (100.0%) | (0.0%) |
|  | max : 2020-07-14 |  |  |  |
|  | range : 6m 14d |  |  |  |
| PM10 | Mean (sd) : 15.7 (7.9) | 61 distinct values | 4747 | 5549 |
|  | min <med <max: |  | (46.1%) | (53.9%) |
|  | 1 <14 <100 |  |  |  |
|  | IQR (CV) : 10 (0.5) |  |  |  |
| PM25 | Mean (sd) : 28.6 (13.7) | 103 distinct values | 10152 | 144 |
|  | min <med <max: |  | (98.6%) | (1.4%) |
|  | 3 <25 <157 |  |  |  |
|  | IQR (CV) : 18 (0.5) |  |  |  |
| NO2 | Mean (sd) : 7.3 (5.8) | 248 distinct values | 6772 | 3524 |
|  | min <med <max: |  | (65.8%) | (34.2%) |
|  | 0.3 <5.6 <48.2 |  |  |  |
|  | IQR (CV) : 5.6 (0.8) |  |  |  |

After matching COVID-19 and WAQI datasets we performed a quality check on the air pollution time series by selecting cities that: (1) Have at least 80% of data coverage after the first case was registered, (2) Have at least 70% of data coverage in the whole period and (3) The estimated precision of the pollution sensor is lesser than 1 µg to PM and NO_2_ and 1 ppm to CO. Since the quality check was employed for each pollutant and location individually, the sets of cities available for each pollutant are not identical.

### 2.2. Granger causality

Granger causality tests were employed to investigate potential relationships between pollution levels (explanatory variables) and the rate of change of COVID-19 new cases (target variable). Granger causality is an econometric method that, when applied on two stationary time series, determines if one can be predicted by past values of the other. Since time series of COVID-19 daily new case are usually “bell” shaped they are highly non-stationary. Therefore, we considered the temporal differences (first derivatives) of the daily new cases time series; i.e., the rate of change of new cases. Then, an Augmented Dickey-Fuller (ADF) unit root test at a critical level of 5% was employed to assert the stationarity of all variables for each city before they were submitted to the Granger causality test.

In the Granger causality test, the target variable was lagged in order to construct a linear autoregressive model. Then, the linear model was expanded to consider lags of the explanatory variable from zero to a maximum lag *n*. An F-test was employed to evaluate if the predictive power of the regression was enhanced by adding the lags of the explanatory variable from zero to *n*. The p-values of the F-test were computed iteratively for *n* ranging from 0 to 20 days. Then, the *n* associated with the most confident p-value was identified for each location. The ADF and Granger causality tests are described in [45]. We employed the implementation available in the Python library *statsmodels*.

### 2.3. Logistic curve fitting

The COVID-19 curve of accumulated cases approximates a logistic-like growth [51, 52]. In a logistic model (Eq. 1), the accumulated cases at a given time (*N* (*t*)) is modeled as a function of the carrying capacity *L* (i.e., the susceptible population), the logistic growth rate *r* (i.e., the steepness of the curve) and the midpoint *t*_0_ which corresponds to the function’s inflection point. The model was fitted for each city using a generalized linear model implementation in the R language *stats* library v3.6.2. Because of its sigmoidal shape, and the “bell” shape of its derivative, the logistic function cannot fit well a time series with more than one “wave” of new cases. To that end, we isolated the first wave of new cases by finding the inflection points (null second derivatives) of the accumulated cases time series; the first wave was defined as the time period before the second inflection point. The time series were smoothed by a 30-day running average before the derivatives were computed. The quality of fitting was quantified by the normalized root mean square error (NRMSE) and Shapiro-Wilk’s test (SW).

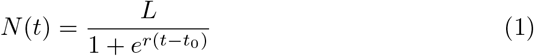

### 2.4. Lag Analysis

The lag analysis is the comparison between two datasets with one being displaced by −*n* samples, where *n* is a positive integer. In this study, we lagged the pollutants concentrations in respect to the new cases time series with *n* ranging from 0 to 30 days. Then, we calculated the time averages of the pollutants concentrations within the first outbreak phase. These averages were linearly regressed against the logistic growth rate. By varying *n*, we explored the time window in which the mean pollution data is best correlated with the logistic model parameters in all cities.

## 3. Results and discussion

### 3.2. Granger Causality

Figure 1 shows the result of the Granger causality test in the counties considered for each pollutant. Counties in green presented F-test p-values below 0.05, while counties in pink are above this threshold. The counties displayed are not the same for all pollutants. This is because, in some locations, the time series either failed the ADF stationarity test or the pollution data failed the quality control.

**Figure 1:**
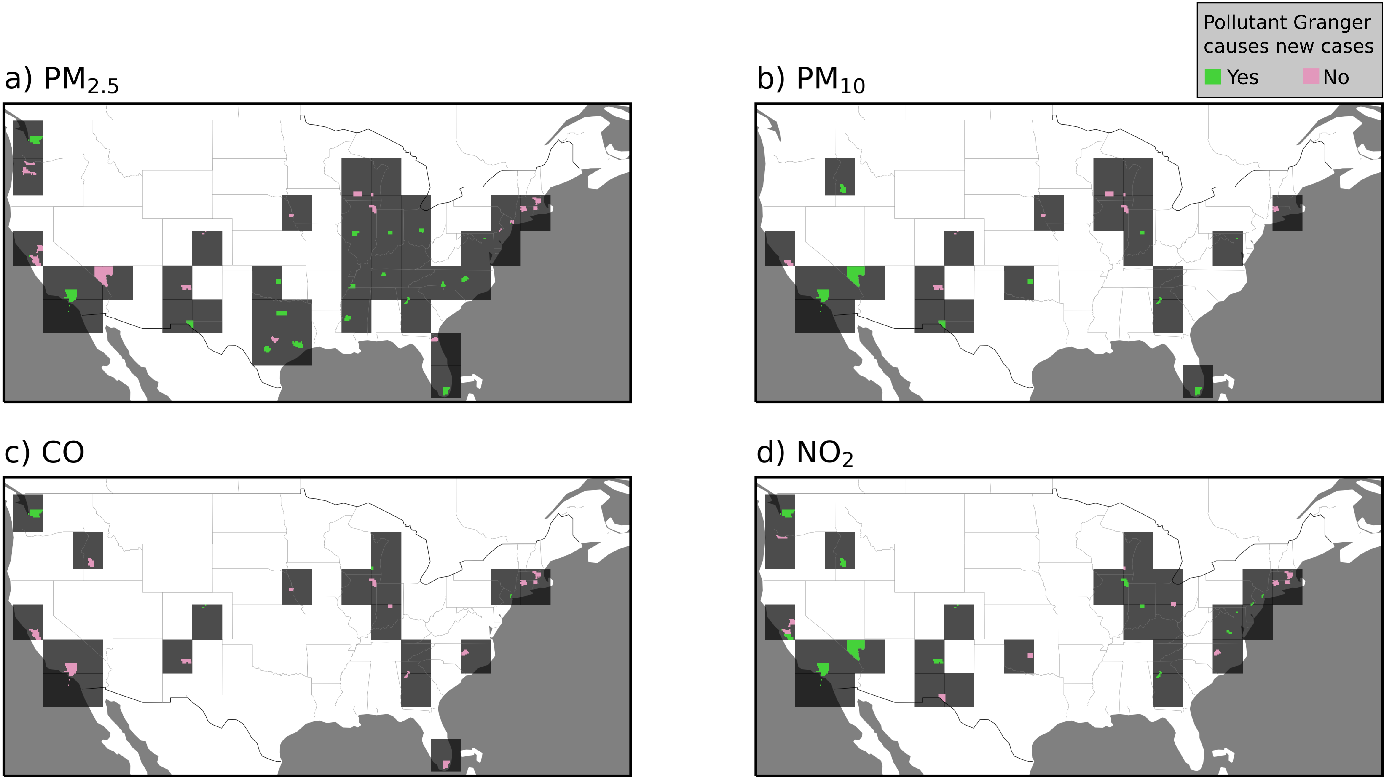
Spatial distribution of the Granger causality test between pollutants concentrations and COVID-19 new cases in USA counties. Counties where the p-values of the Granger F-test are green when below 0.05 and pink when above. Dark gray squares were plotted around colored counties to facilitate their visualization.

It is readily visible that PM_2.5_ (Fig. 1a) Granger causes new COVID-19 cases in a substantial number of locations. This is also the case for PM_10_ (Fig. 1b) and NO_2_ (Fig. 1d); however, less locations are available for these pollutants. CO, on the other hand, visibly failed the F-test in most locations. The spatial distributions of locations where pollutants Granger cause new cases do not show any clear pattern, such as a preferential latitude. This is evidence against possible confounding effects, such as weather conditions or other regional particularities.

The boxplot in Figure 2a shows the distribution of p-values of the Granger F-test between the pollutants and COVID-19 new cases considering the locations shown in Figure 1. PM_2.5_ shows a significant Granger causal link to the rate of change of COVID-19 new cases in 17 of 44 locations and presented the lowest p-value mean among the pollutants. NO_2_ presented a lower median, however only 7 out of 28 locations presented p-value under the threshold of 0.05. PM_10_ mean and median p-value are higher than PM_2.5_ and PM_10_; in this case, 8 out of 20 locations passed the significance test. CO presented the highest p-value mean and median; only in 4 out of 21 locations a significant causality link was found. Figure 2b shows the distribution of lag windows for locations where there is a significant causal link between pollution and COVID-19 new cases. This allows us to verify that the SARS-CoV-2 incubation period is within the range of the lag windows encountered by the Granger analysis, sustaining the plausibility of the hypothesis.

**Figure 2:**
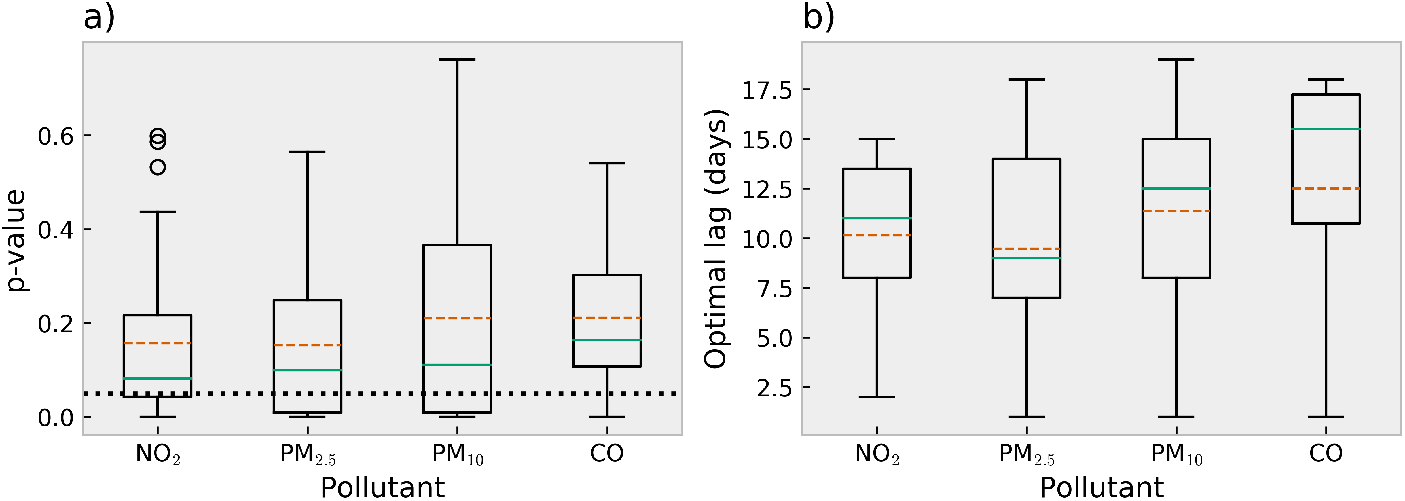
a) Boxplot of p-values of the Granger F-test between pollutants concentrations and COVID-19 new cases; b) boxplot of the optimal lag window size only considering locations where the p-values of the Granger F-test are below 0.05. Medians are teal solid lines and means are orange dashed lines. In a), the black dotted line identifies p-value = 0.05.

### 3.2. Logistic Model

For each city available for the pollutants we fitted a logistic curve to the accumulated cases data. The period selected in each city starts on the day where sixth case was recorded to the first minimum of the COVID-19 new cases (first phase of outbreak). The first minimum was obtained after smoothing the time series with 30-day moving averages. This was done in order to isolate the “first phase” or “first wave” of the epidemic because the logistic model supports a single sigmoid curve. The average NRMSE of the fits is 0.0273*±*0.0096 %. The average SW p-value for the fitted models is 0.0141*±*0.0231. The NRMSE lesser than 0.05 indicate a good overall fit. SW p-values lesser than 0.05 indicate normally distributed residuals without biases. Both indicates that the logistic model is well fitted to the curve of COVID-19 accumulated cases. Thus, the model parameters can be used to represent aspects of the observed data.

### 3.2. Lag Analysis

Pearsons’s correlation coefficient and their p-values (for the hypothesis of null correlation) were calculated between the average pollutant concentration and the logistic growth rate (Figure 3) in order to assert the relationship between the pollutants and the outbreaks growth rate. PM_2.5_ presents higher and significant correlations from lags 0 to 18 days. CO presents significant correlations at all lags; its correlations are higher in later lags, in special after 26 days. Since CO is a good tracer to fossil fuel usage, large temporal shifts places the pollutant data outside the quarantine range. Thus, the rise of correlation at later lags is expected as it enhances the representation of the overall social mobility before the outbreak in the cities. This behaviour is also observed in PM_10_ and NO_2_, although the latter do not present significant correlations.

**Figure 3:**
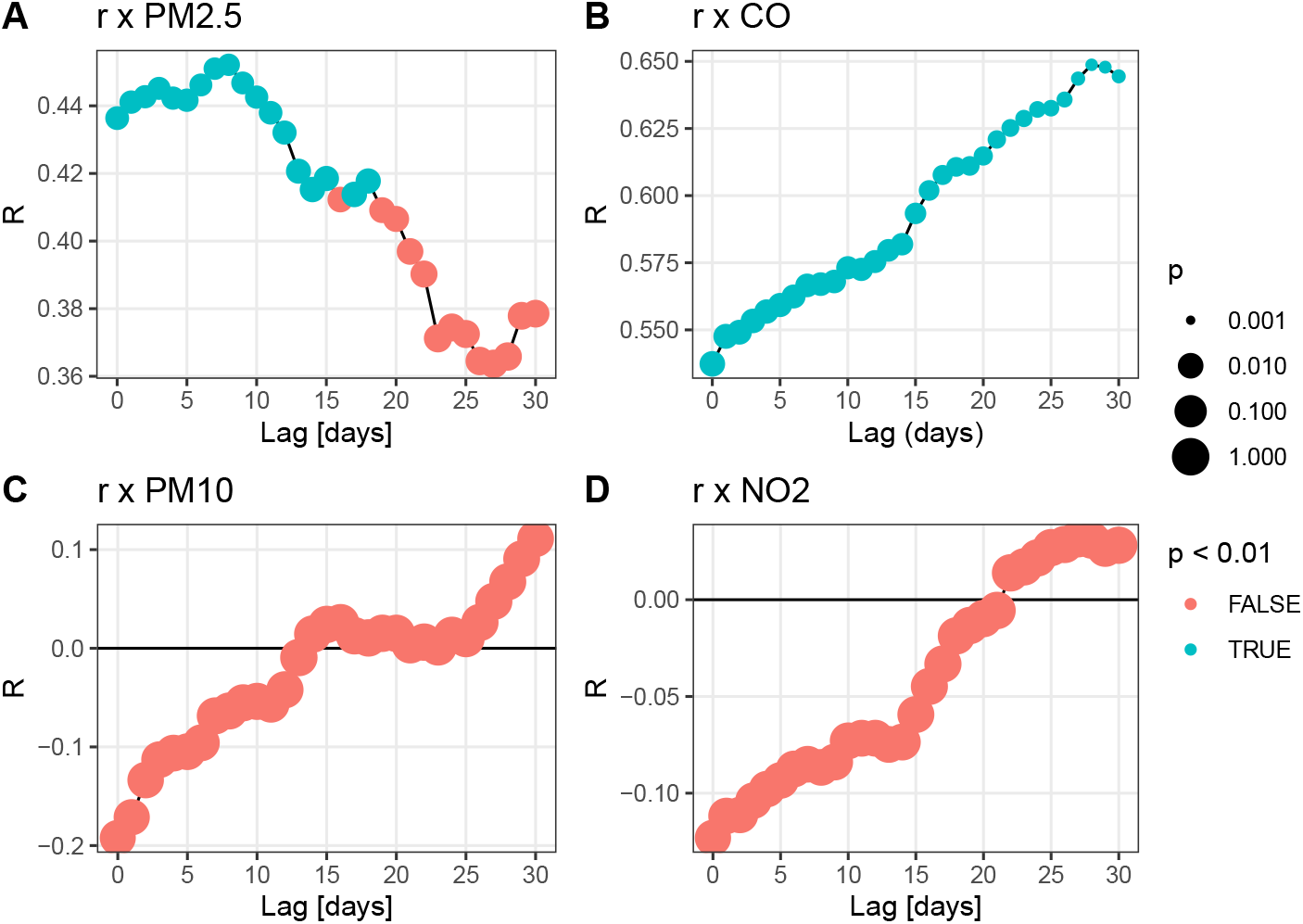
Pearson’s correlation between the logistic growth rate and the pollutants for all cities for lags between 0 to 30 days. p-values for the zero zero correlation null-hypothesis are represented by the point size. p-values lesser than 0.01 are in red. The opposite is in blue.

PM_2.5_ presents higher lags in a period that matches the SARS-CoV-2 incubation time. It support the Granger analysis about the short-term role of PM_2.5_. However the lag analysis points out a significant impact in the outbreak development, assigning a major role to PM_2.5_.

### 3.4. Quantifying the general impact of the pollutant sources

We used linear regressions to quantify the impact of PM on the spread of COVID-19 characterized by the logistic growth rate (Figure 4). Other regressions may be more appropriated, but they require justification based on previous knowledge of the mechanisms behind the PM/COVID-19 interaction. Therefore, the linear regression was employed as a naive method to determine the general tendency of these relationships.

**Figure 4:**
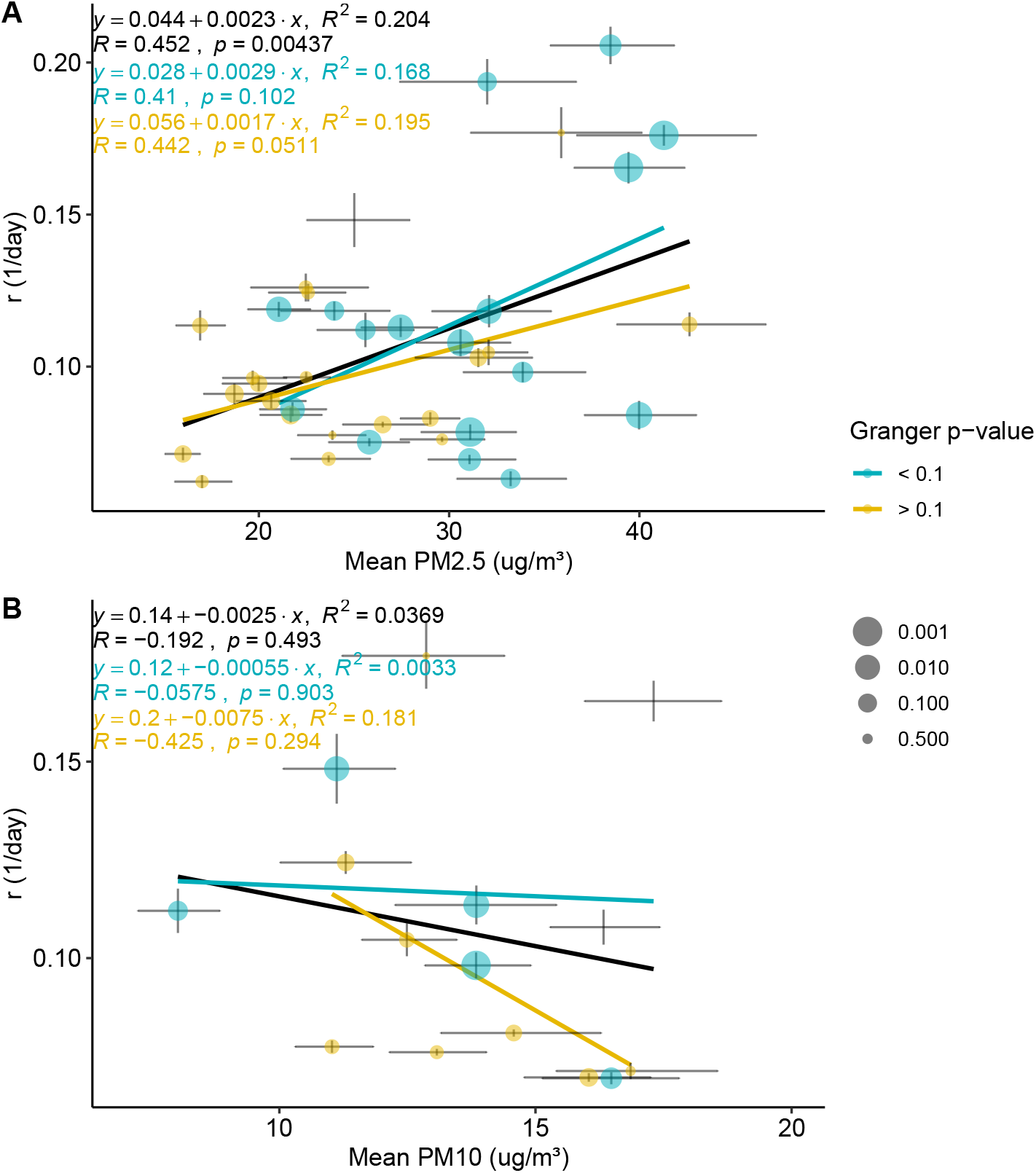
Logistic growth rate of COVID-19 accumulated cases (r) as a function of mean pollutants concentrations. The average pollutants concentrations are in the best correlated lag calculated in the Lag Analysis section (8, and 0 days to PM_2.5_ and PM_10_ respectively). The linear regression with all cities is represented by the black line. Parameters of this linear regression are in each plot. Cities where the pollutant Granger causes new daily cases (described in Granger Causality section) are presented in blue, the rest in yellow. The size of points represents the p-value of the Granger test, greater sizes represent smaller p-values. Linear regression to the blue and yellow groups of points are presented by the blue and yellow lines. A version with the cities labeled in the plots is available in the Supplementary Material. The horizontal error bars were calculated using bootstrap with 1000 interactions. The vertical error bars were obtained from the logistic fit.15

In Figure 4A the general impact of PM_2.5_ in the outbreak is clear. We note, for example, that an increase in PM from 20 to 40 µg *m*^−3^ rises the logistic growth rate from 0.09 to 0.136 1/day, corresponding to a 54% increase. If we only consider cities where the PM_2.5_ Granger causes the COVID-19 new cases, the same PM_2.5_ increase rises the logistic growth rate from 0.086 to 0.144 1/day, corresponding to a 67% increase. Considering cities where the Granger test fails, the same PM increase only rises the growth rate from 0.09 to 0.124 1/day (i.e., 38%). A similar behaviour occurs with the 0.1 and 0.9 quantiles of the PM_2.5_ (see supplementary material). *PM*_10_ (Figure 4B) shows no significant correlation with the logistic growth rate.

The interpretation of PM_2.5_/COVID-19 statistical relationships is not straight-forward. We can describe at least three potential mechanisms underpinning these relationships: (1) long-term PM_2.5_ exposure increases population susceptibility; (2) PM_2.5_ indicates social mobility and (3) PM_2.5_ is a viral airborne transport facilitator. Mechanisms 1 and 2 are confounding factors to Mechanism 3, so we will discuss them individually.

Mechanism 1 is expected to be present in all regressions in Figure 4A, as cities with higher mean PM_2.5_ in the studied period are likely to have an atmospheric polluted history. To separate the more instantaneous role of PM_2.5_ (Mechanism 3), we can consider the difference between the regression using cities where PM_2.5_ Granger causes new daily cases (blue line in Fig. 4A) and cities where the Granger test fails (yellow line). The higher slope of the blue regression indicates that Mechanism 3 is discernible from the confounding factor posed by Mechanism 1. Mechanism 2, however, does not seem to be an important confounding factor for Mechanism 3 as the lagged correlations between PM_10_ and NO_2_ are inconclusive and CO only passes the Granger test in four locations. Using the difference of the blue and yellow linear regressions in Fig. 4A, we calculate a rise of the logistic growth of 0.012 1/day based on an increase from 25 to 35 µg *m*^−3^ of PM_2.5_. For instance, the growth rate of accumulated cases in Boston of 0.096 1/day and PM_2.5_ averages at 25 µg *m*^−3^. An increase of 10 µg *m*^−3^ of PM_2.5_ rises its growth rate by approximately 12.5%.

## 4. Conclusion

In this study we explored the short-term role of the particulate matter in the COVID-19 outbreak in USA cities. We applied the Granger’s causality tests, lag analysis and logistic modelling to investigate the statistical links between the spread of COVID-19 and pollution data. The comparison between PM and the other pollutants allowed us to isolate PM_2.5_ from confounding factors and estimate its contribution to airborne viral transportation. The findings support the viral transport hypothesis, i.e., virus can associate with the pre-existent particulate matter in the air synergically. We conclude that PM_2.5_ plays a small, yet discernible, role in the COVID-19 transmission.

The USA presents diverse geographic, climatic and political scenarios. This suggests that the conclusions presented here could be potentially be generalized to other countries. Increasingly abundant COVID-19 data worldwide will facilitate future studies to explore these interactions in a global scale. Broadly, we hope to rise the interest of the scientific community as well as the awareness of the general public and decision makers to the potential synergy between viral transmission and air pollution.

Considering that we are still with increasing mortality rate in most countries, any efforts to decrease the transmissibility of the COVID can be summed up with all the others to save some lives.

## Data Availability

All datasets included in this article are open and publicly available

https://github.com/lokamigauti/COVID_Article

## 5. Acknowledgements

We thank the Brazilian agencies CAPES, CNPq and FAPESP for funding this study. We also thank all the essential workers that are risking theirs lives in favor of society.

