## Supplemental tables for "Exploring the short-term role of particulate matter in the COVID-19 outbreak in USA cities"

February 2021

This supporting information has 5 pages including Tables S1-S4.

| | County | State | p-value |
|---|---|---|---|
| 1 | Fort Worth | Texas | 0.00 |
| 2 | Jacksonville | Florida | 0.00 |
| 3 | Nashville | Tennessee | 0.00 |
| 4 | San Antonio | Texas | 0.00 |
| 5 | Oklahoma City | Oklahoma | 0.00 |
| 6 | Raleigh | North Carolina | 0.00 |
| 7 | Miami | Florida | 0.00 |
| 8 | Jackson | Mississippi | 0.00 |
| 9 | Springfield | Illinois | 0.01 |
| 10 | Memphis | Tennessee | 0.01 |
| 11 | Staten Island | New York | 0.01 |
| 12 | Los Angeles | California | 0.01 |
| 13 | Seattle | Washington | 0.01 |
| 14 | Columbus | Ohio | 0.01 |
| 15 | Indianapolis | Indiana | 0.02 |
| 16 | Houston | Texas | 0.04 |
| 17 | Atlanta | Georgia | 0.05 |
| 18 | Baltimore | Maryland | 0.06 |
| 19 | Dallas | Texas | 0.07 |
| 20 | San Francisco | California | 0.07 |
| 21 | Charlotte | North Carolina | 0.09 |
| 22 | El Paso | Texas | 0.10 |
| 23 | Providence | Rhode Island | 0.10 |
| 24 | Portland | Oregon | 0.10 |
| 25 | Brooklyn | New York | 0.13 |
| 26 | Omaha | Nebraska | 0.14 |
| 27 | Little Rock | Arkansas | 0.15 |
| 28 | Albuquerque | New Mexico | 0.15 |
| 29 | Manhattan | New York | 0.15 |
| 30 | Philadelphia | Pennsylvania | 0.18 |
| 31 | Sacramento | California | 0.18 |
| 32 | Las Vegas | Nevada | 0.22 |
| 33 | Austin | Texas | 0.23 |
| 34 | Oakland | California | 0.31 |
| 35 | Denver | Colorado | 0.31 |
| 36 | San Jose | California | 0.34 |
| 37 | Salem | Oregon | 0.36 |
| 38 | Queens | New York | 0.36 |
| 39 | Detroit | Michigan | 0.38 |
| 40 | Boston | Massachusetts | 0.39 |
| 41 | Milwaukee | Wisconsin | 0.40 |
| 42 | Chicago | Illinois | 0.42 |
| 43 | Hartford | Connecticut | 0.56 |
| 44 | Madison | Wisconsin | 0.56 |

Table 1: Results of Granger Analysis for PM25.

| | County | State | p-value |
|---|---|---|---|
| 1 | Miami | Florida | 0.00 |
| 2 | Jacksonville | Florida | 0.00 |
| 3 | Oklahoma City | Oklahoma | 0.00 |
| 4 | Atlanta | Georgia | 0.00 |
| 5 | Las Vegas | Nevada | 0.01 |
| 6 | Boise | Idaho | 0.01 |
| 7 | El Paso | Texas | 0.02 |
| 8 | Indianapolis | Indiana | 0.05 |
| 9 | Los Angeles | California | 0.08 |
| 10 | Baltimore | Maryland | 0.09 |
| 11 | Denver | Colorado | 0.13 |
| 12 | San Jose | California | 0.19 |
| 13 | Omaha | Nebraska | 0.23 |
| 14 | Detroit | Michigan | 0.30 |
| 15 | Hartford | Connecticut | 0.35 |
| 16 | Chicago | Illinois | 0.42 |
| 17 | Honolulu | Hawaii | 0.43 |
| 18 | Milwaukee | Wisconsin | 0.46 |
| 19 | Albuquerque | New Mexico | 0.68 |
| 20 | Madison | Wisconsin | 0.76 |

Table 2: Results of Granger Analysis for PM10.

| | County | State | p-value |
|---|---|---|---|
| 1 | Denver | Colorado | 0.00 |
| 2 | Staten Island | New York | 0.00 |
| 3 | Seattle | Washington | 0.01 |
| 4 | Milwaukee | Wisconsin | 0.02 |
| 5 | Manhattan | New York | 0.07 |
| 6 | San Jose | California | 0.11 |
| 7 | Raleigh | North Carolina | 0.11 |
| 8 | Albuquerque | New Mexico | 0.13 |
| 9 | Boise | Idaho | 0.15 |
| 10 | Oakland | California | 0.15 |
| 11 | Indianapolis | Indiana | 0.16 |
| 12 | Atlanta | Georgia | 0.21 |
| 13 | Chicago | Illinois | 0.22 |
| 14 | Providence | Rhode Island | 0.22 |
| 15 | Los Angeles | California | 0.26 |
| 16 | Omaha | Nebraska | 0.30 |
| 17 | Hartford | Connecticut | 0.35 |
| 18 | San Francisco | California | 0.42 |
| 19 | Jacksonville | Florida | 0.49 |
| 20 | Miami | Florida | 0.50 |
| 21 | Boston | Massachusetts | 0.54 |

Table 3: Results of Granger Analysis for CO.

| | County | State | p-value |
|---|---|---|---|
| 1 | Denver | Colorado | 0.00 |
| 2 | Seattle | Washington | 0.00 |
| 3 | Boise | Idaho | 0.00 |
| 4 | Manhattan | New York | 0.01 |
| 5 | Baltimore | Maryland | 0.02 |
| 6 | Indianapolis | Indiana | 0.02 |
| 7 | San Francisco | California | 0.02 |
| 8 | Staten Island | New York | 0.05 |
| 9 | Philadelphia | Pennsylvania | 0.05 |
| 10 | Chicago | Illinois | 0.06 |
| 11 | Albuquerque | New Mexico | 0.07 |
| 12 | San Jose | California | 0.07 |
| 13 | Atlanta | Georgia | 0.08 |
| 14 | Los Angeles | California | 0.08 |
| 15 | Las Vegas | Nevada | 0.08 |
| 16 | Richmond | Virginia | 0.09 |
| 17 | Hartford | Connecticut | 0.11 |
| 18 | El Paso | Texas | 0.12 |
| 19 | Providence | Rhode Island | 0.12 |
| 20 | Boston | Massachusetts | 0.14 |
| 21 | Sacramento | California | 0.20 |
| 22 | Milwaukee | Wisconsin | 0.27 |
| 23 | Portland | Oregon | 0.27 |
| 24 | Columbus | Ohio | 0.30 |
| 25 | Jacksonville | Florida | 0.44 |
| 26 | Raleigh | North Carolina | 0.53 |
| 27 | Oakland | California | 0.59 |
| 28 | Oklahoma City | Oklahoma | 0.60 |

Table 4: Results of Granger Analysis for NO2.
